# Novel indicator of change in COVID-19 spread status

**DOI:** 10.1101/2020.04.25.20080200

**Authors:** Takashi Nakano, Yoichi Ikeda

## Abstract

In the fight against the pandemic of COVID-19, it is important to be quick to detect changes in the rate of spread and to be precise to predict the future. We have succeeded in formulating a new indicator based on daily number of infected people from publicly available data, which enables us to do the both. We show the validity of the indicator by demonstrating that a universal analysis of current status of COVID-19 spreading over countries can predict effects of measures such as the blockage of cities and social distancing, signs of new spread, and possible regional dependence in the formation of herd immunity.

## Introduction

Global spread of COVID-19 has resulted in significant human and economic losses worldwide. In order to prevent the spread of infection, it is necessary to restrict social activities by policies such as the blockade of cities and the prohibition of assembly. For the effective implementation of these policies, it is necessary to ascertain the status of spread and to estimate the trend of spread accurately. However, it is often difficult to grasp the severity of spread and estimate the trend by using model calculations because the implementation criteria of PCR testing vary from country to country and a high level of expertise is required to adjust model parameters according to the circumstances of each country. The threat of COVID-19 has spread over countries which do not have the high-level computing resources, and the development of means to ascertain the status of spread accurately without relying on specific models has become an urgent issue.

## Indicator *K* to analyze the transition of COVID-19 spread

In this study, we introduce a new indicator called the *K* value defined by *K*(*d*) *=* 1 − *N*(*d* − 7)*/N*(*d*), where *d* is the number of days from the reference date, and *N*(*d*) and *N*(*d* − 7) are the total number of infected people on days *d* and (*d* − 7), respectively. Since *N*(*d*) is greater than *N*(*d* − 7) during the period from the initiation of spread to convergence, *K* takes a value between 0 and 1.

Without loss of generality, the daily evolution of *N*(*d*) can be expressed with a time dependent exponential factor *a*(*d*) as *N*(*d* + 1) = exp(*a*(*d*)) *N*(*d*). Our assumption is that *a*(*d*) can be expressed by a geometric series with a constant dumping factor *k*, namely, *a*(*d* + 1) = *ka*(*d*). The simulation study under this assumption found that *K* can be approximated by a first order linear function of *d* in a wide range (0.25 < *K* < 0.9) and the input value of *k* can be reproduced by *k* = 1 + 2.88*K′*, where *K′* is a slope of a straight line obtained by the fit (see Supplementary materials). The validity of the assumption has been checked and confirmed by analyzing the existing data (*1–3*) as demonstrated in the following paragraphs.

The quotient of |*K*/*K′*| gives a good estimation for a time period for COVID-19 spread to converge, and an upward change of the *K* trajectory indicates a new outbreak. By updating the *K* value in real time using daily input data, we can identify the current status of spread, estimate the future status of spread, and detect signs of new spread at an early stage.

## Analyses of China and USA using the *K* indicator

The analysis using *K* was first applied to China, which was less susceptible to the impact of other countries, because the spread began ahead of other countries. The *K* value was calculated using data from January 26 onward, but the data before February 12 were uniformly multiplied by 1.27 in order to correct a sharp increase in the number of infected people artificially caused by the change of certification criteria for SARS-CoV-2 infections in Hubei Province, China on February 13. As shown in Figure 1, the *K* values is closely approximated by a straight line with a slope (*K′*) of −0.0402±0.0008/*d*. The linearity of *K* is also prominent in the United States. After a high *K* level period indicating successive infectious explosions from mid to late March, the *K* has continued to decline with a uniform rate of *K*′ *=* −0.0237 ± 0.0003/*d* in the fitting region of 0.25 < *K* < 0.90. In a region of *K* < 0.25, transition of *K* can be estimated with a constant dumping factor *k* calculated from *K′*. Comparison between the data points and the model estimations tells us that the pace of convergence became higher in China in the terminating phase, while it was significantly reduced in USA, indicating that the spread may have turned to expand again. Since the transition of the *K* value in New York state remains normal, there may have been outbreaks in the states where the COVID-19 spread was less serious.

**Figure 1:**
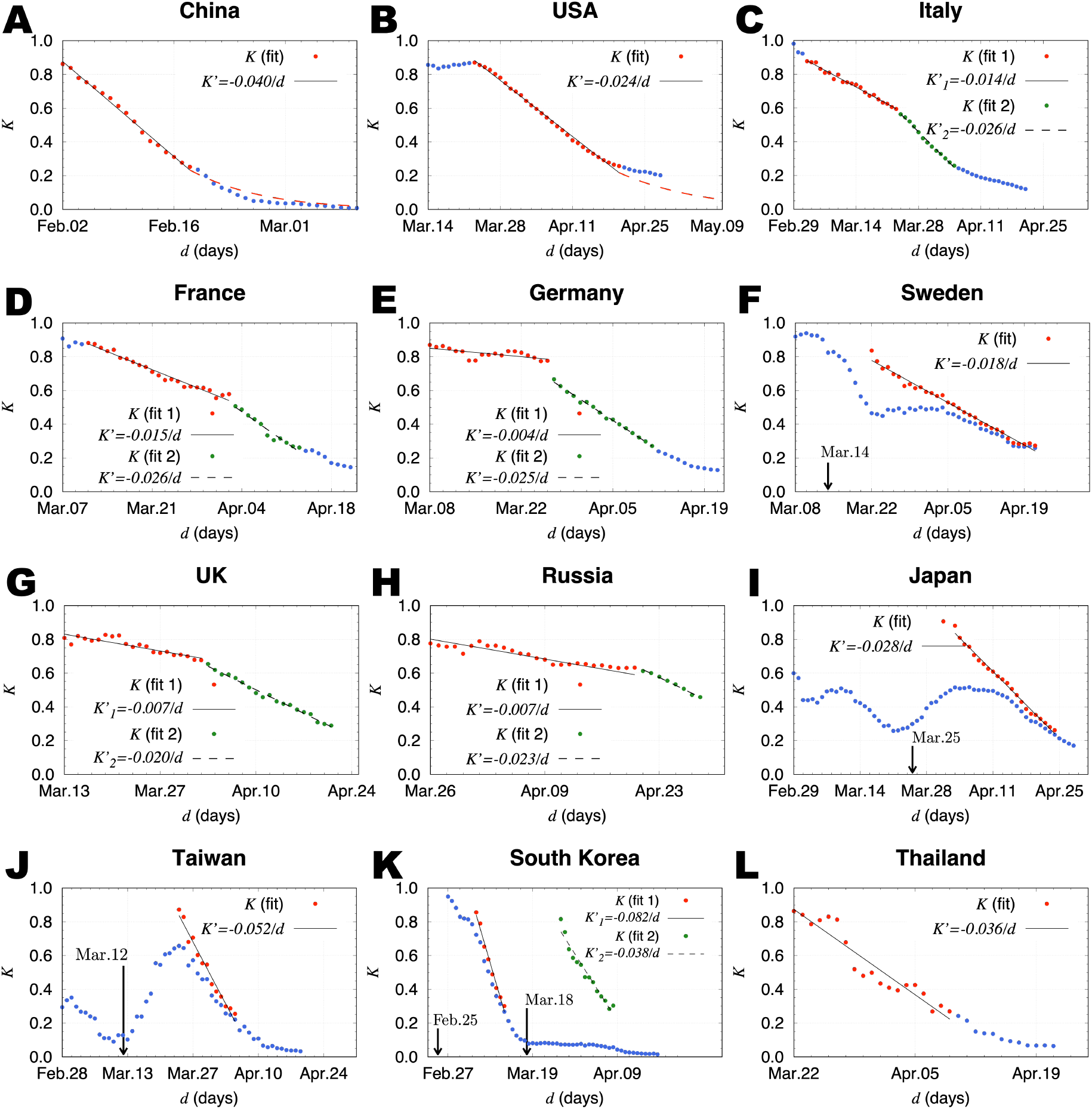
Transition of the K values from February to April, 2020. (**A**), The *K* values of China obtained from the daily total number of infected people. The slope *K′* was obtained by a linear fit in the range of 0.25 < *K* < 0.9. The data points used for the fit are indicated by red points. The solid line is the fit result. The dashed curve is estimated with the constant dumping factor *k* calculated from *K′*. (**B**), The *K* values of USA. (**C**), The *K* values of Italy. The first and second *K′* values were obtained by linear fits using red and green points, respectively. The solid line is the result of the first fit. The dashed line is the result of the second fit. (**D**), The *K* values of France. (**E**), The *K* values of Germany. (**F**), The *K* values of Sweden. The total number of infected people were counted from the reference date set on March 14. (**G**), The *K* values of UK. (**H**), The *K* values of Russia. (**I**), The *K* values of Japan. The reference date was set on March 25. (**J**), The *K* values of Taiwan_8_ The reference date was set on March 12. (**K**), The *K* values of South Korea. The reference dates were set on February 25 and March 18 for the first and second fits, respectively (**L**), The *K* values of Thailand.

In order to understand the change of *K*, the analysis of the data for the United States is performed using the SI epidemic model (*4*) (see also Supplementary materials). It is found that we need to take into account at least the four independent sources of the infection to reproduce the data. We find the peak positions of the maximum momentum of the infection with respect to each source as March 26, April 4, 14 and 24 (Fig. 2 **A**). The first peak position clearly corresponds to the date of the change in the slope of the *K*. This shows that the *K* starts to decrease when the first peak-out takes place. The other sources contribute to keep the *K* linear as superposition.

**Figure 2:**
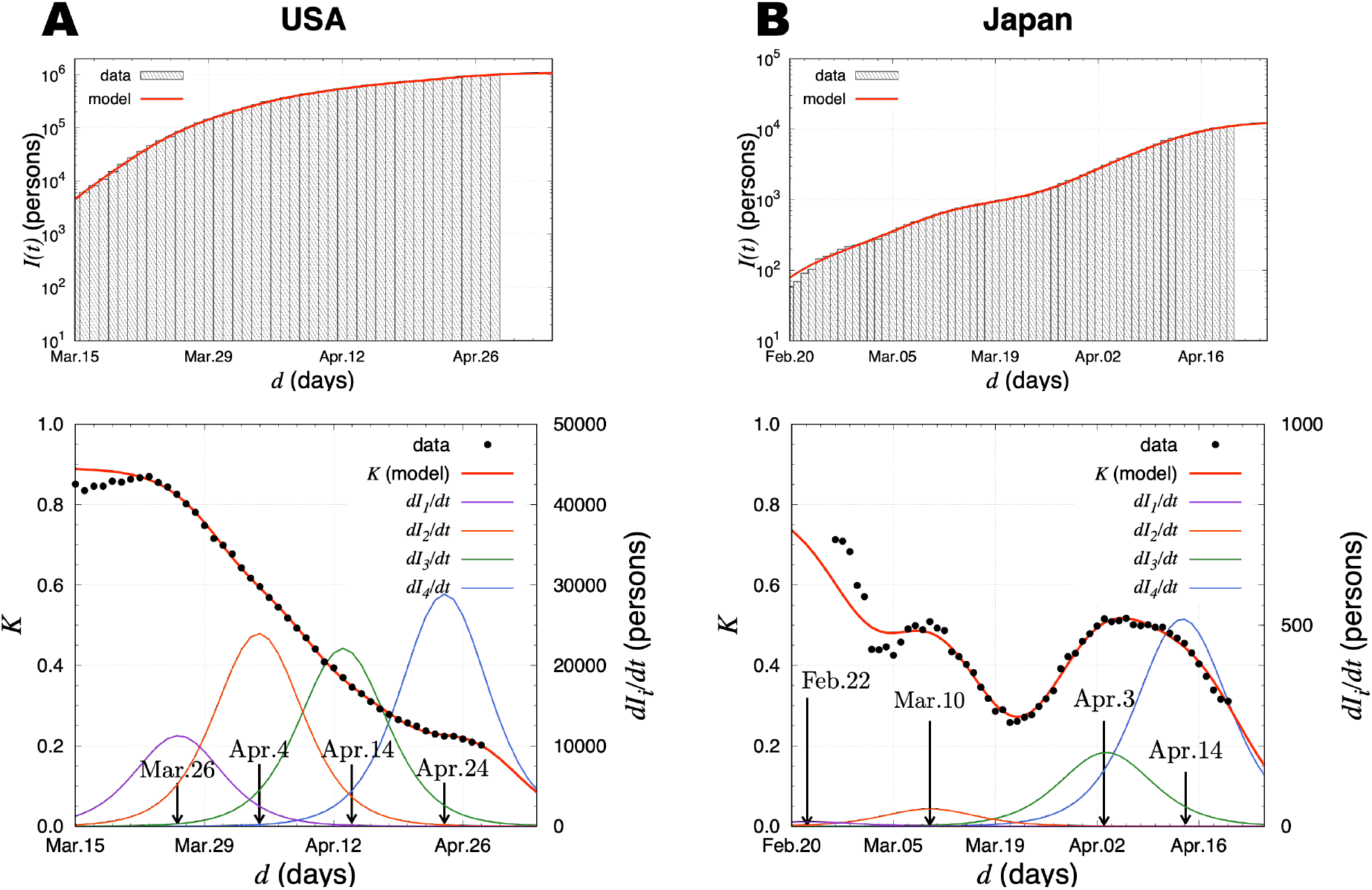
The model results of total number of infected people and the *K* value in the United States and Japan. (**A**), The results for the United States. The fit results are given in the top figure. The *K* value is represented together with the daily new cases *dI_i_/dt* calculated in the SI model in the bottom. The peak positions 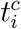 caused by the source *i* (*i* = 1, 2, 3, 4) are shown by the vertical allows. (**B**), The results for Japan. Same as (A) for the case of Japan.

## Spread of COVID-19 and countermeasures in European countries

In Italy, where COVID-19 started to spread first in Europe, *K′* was −0.0142 ± 0.0004/*d* from March 1 to 23 indicating a slow pace of convergence, but the pace was improved after March 24 resulting in *K′* = −0.0263 ± 0.0006/*d*. This improvement is most likely due to the containment policy implemented in early March, including the blockade of cities. In France, where the spread started after about 10 days from Italy, the transition of *K* has similar tendencies. The *K′* was −0.0152 ± 0.0005/*d* for three weeks in the early stages, and it was improved to −0.0265 ± 0.0019/*d*. Countries’ policies changed the *K′* values in both Germany and Sweden. On March 16, Germany began measures to prevent the spread of infection, including closure of most retail stores except grocery stores and pharmacies, restriction of operation hours of restaurants, and prohibition of assembly at religious facilities. This resulted in a steep *K* slope (*K′* = −0.0252 ± 0.0005/*d*). Sweden, on the other hand, sought to acquire herd immunity and took relatively mild measures resulting in a moderate gradient (*K′* = −0.0179 ± 0.0004/*d*) lasting more than a month. UK took similar measures in the early stage of the spread resulting in infection explosion with *K′* = −0.0071 ± 0.0008/*d*. It was tamed down (*K′* = −0.0199 ± 0.0005/*d*) after they introduced the strict policy. In Russia, the *K* value stayed above 0.6 with a slope of *K′* = −0.0067± 0.0005/*d* until April 20, but it started to decline with a slope of *K′* = −0.0234 ± 0.0010/*d*, indicating that a catastrophic situation was avoided.

It is quite important to minimize the period of the early stages with *K′* ~ −0.007/*d*. One week delay of implementation of countermeasures will double the total number of infected people.

## Transition of COVID-19 spread in Asian countries

In Asian countries close to China, after the first wave originated in China, and the subsequent spread in synchronized with the worldwide spread can be observed as the upward change in *K* trajectories. In order to accurately estimate the change in the status of the subsequent spread from the *K* and *K′* values, it is necessary to set a reference date for counting the total number of infected people at the rise of the second wave. We obtained *K′* = −0.0283 ± 0.0006/*d* in Japan by setting the reference date on March 25. The slope is milder than those of Taiwan (*K′* = −0.0524 ± 0.0026/*d*) and South Korea (*K′* = −0.0820 ± 0.0042/*d* and *K′* = −0.0378 ± 0.0024/*d*), reflecting the difference in the strictness and efficiency of countermeasures. However, it is steeper than those of European countries with more strict social restrictions than Japan. The relatively high absolute *K′* values even in the early stages are common in many Asian countries. For example, the *K′* value of Thailand is *K′* = −0.0361 ± 0.0028/*d*. This characteristic high |*K′*| value suggests that the herd immunity may be formed more quickly in these countries than European countries (*5*, *6*). An antibody testing survey in Japan and other Asian countries with relatively mild measures will reveal the truth. Incidentally, the difference between the two *K′* values in South Korea may be attributed to the fact that most of the infection routes were clear in the former, whereas the route was often unknown in the latter.

To reproduce Japan data (*3*) with the SI epidemic model, which has two peaks in the *K* from March to April, it is necessary to introduce four sources of the spread. The corresponding peak positions are obtained as February 22, March 10, April 3 and 14. In the actual data, the *K* starts to decrease on March 12 in the first peak and on April 11 in the second one. The peak positions in the model agree with these date (Fig. 2 **B**). Moreover, the third peak position in the model coincides with the date when the *K* saturates at 0.5.

Comparing with the model analysis of the United States, it is worth mentioning that the parameter, which controls how infections spread, is smaller in Japan than that in the United States. Nevertheless, the steep gradient of the *K* is observed in Japan. This observation is understood by the deference of the number of the sources of the infection contributing to the *K* in the both countries. In Japan, the first two and last two sources are well separated in time, so that only the last two affect the decrease of the *K* as superposition. While, in the United States, all four sources contribute to it. The results show the *K* plays a crucial role to understand how the infection spreads.

## Conclusion

As the spread of COVID-19 worldwide progresses, it is important not only to protect human lives, but also to minimize social losses due to economic paralysis by detecting signs of spread at an early stage and predicting future trends accurately. We have demonstrated that the value of *K* and its slope of *K′* are crucial for understanding the spread status of COVID-19 for these purposes. Thanks to linear dependence on the elapsed days and stability due to one-week interval for the calculation, extrapolation of the *K* trajectory and prediction of the future trend of COVID-19 are easy. Analyses with the *K* and *K′* values will help us to implement appropriate measures in a timely manner and evaluate their effects. Moreover, since the *K′* is related with a fundamental factor *k*, a global antibody testing survey together with a systematic study of *K′* may reveal the underlying reasons for regional differences in infection rate and mortality of COVID-19 between Europe and Asia. Also, as evidenced by the comparison with the SI model calculations, the linearity of the *K* value is not trivial but is most likely to be caused by several consecutive infectious explosions. Focusing on the change in the value of *K* will help to improve and refine epidemiological models of infectious diseases with the same tendency as COVID-19.

## Data Availability

We collected data from publicly available data sources:
https://ourworldindata.org/coronavirus-source-data
https://web.sapmed.ac.jp/canmol/coronavirus/index.html
The COVID Tracking Project,
https://covidtracking.com/data/us-daily
NIPPON TELEVISION NETWORK CORPORATION, COVID-19 special site.
https://www.news24.jp/archives/corona_map/index2.html

## Acknowledgments

We thank Prof. Y. Kaneda (Vice President of Osaka University), Prof. T. Yoshimori (Graduate School of Frontier Bioscience, Osaka University), and Mr. S. Shimasaki (Embassy of Japan in the United States of America) for helpful discussions.

## Supplementary materials

Materials and Methods

Figs. S1 and S2

### Materials and Methods

We assumed that a convergent series of the total number of infected people (*N*(*d*)) is given by

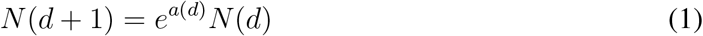

and

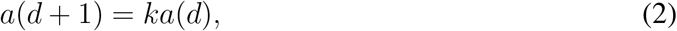

where *k* is a constant (*k* < 1). Then *K* values for *d* > 7 were calculated by *K*(*d*) = 1 − *N*(*d* − 7)/*N*(*d*) for the cases with *k* = 0.890, 0.895, 0.900, 0.905, 0.910, 0.915, 0.920, 0.925, 0.930, 0.935, 0.940 and 0.950 (Fig. S1). The fitting region of 0.25 < *K* < 0.9 with a linear function was determined so that the maximum deviation of data points from the straight line is less than 11% of the data value. The relation between the slope of the straight line *K′* and *k* was examined to obtain a linear relation of *k* = 1 + 2.88*K′* (Fig. S1). Note that the relation does not depend on *a*(0) as far as the first *K* value is larger than 0.9. We set *a*(0) = 0.5 for the all cases. For application of real-data analyses, once *k* is known from *K′*, the daily evolution of *N*(*d*) can be recursively calculated from the equations (1) and (2) as shown in Fig. S2.

In order to understand the behavior of the *K*, we analyze the public data of COVID-19 in the United States and Japan employing the SI epidemic model (*4*), which consists of the infectives(*I*) and susceptibles(*S* = *N* − *I*) with *N* being the final number of infectives. In the both countries, several changes in the slope of the *K* are found, it is natural to introduce several sources of COVID-19: The *K* is a monotonically decreasing function if a single source is taken into account in the SI model. The infectives with respect to each source *i* is described as

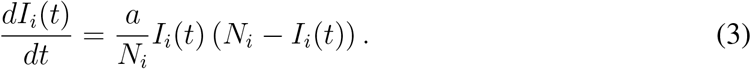

The model parameters *a* and *N_i_* control the spreading speed of COVID-19 and the final number of infectives caused by the source *i*, respectively. The analytic solution for *I_i_* is found as 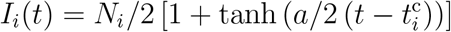 with 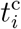 denoting the peak time of the infection spreading. The total number of infected people in a country at time t are then obtained as 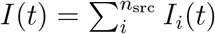 and finally reach 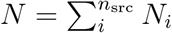. The optimal solution of the parameter set 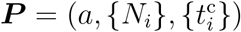 together with the number of sources *n*_src_ is found by minimizing the weighted mean-square deviation *L*(***P***) = ∑*_d∊D_* (*I*(*d*) − *N*(*d*))^2^ /*N*(*d*) in the fit range ***D***. We choose ***D*** from February 23 to April 20 (58 days) for Japan and March 15 to April 28 (45 days) for the United States, respectively. We find the optimal number of sources for both countries as *n*_src_ = 4, and the resulting parameters are given by (*a*, *N*_1_, *N*_2_, *N*_3_, *N*_4_, 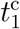, 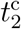, 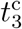, 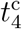) = (0.32, 142k, 303k, 279k, 365k, 03/26, 04/04, 04/14, 04/24) for the United States and (0.24, 0.2k, 0.7k, 3.1k, 8.7k, 02/22, 03/10, 04/03, 04/14) for Japan, respectively. Equipped with the parameters above, the number of daily new cases *dI_i_*/*dt* and the *K* value are presented in Fig. 2.

**Fig. S1:**
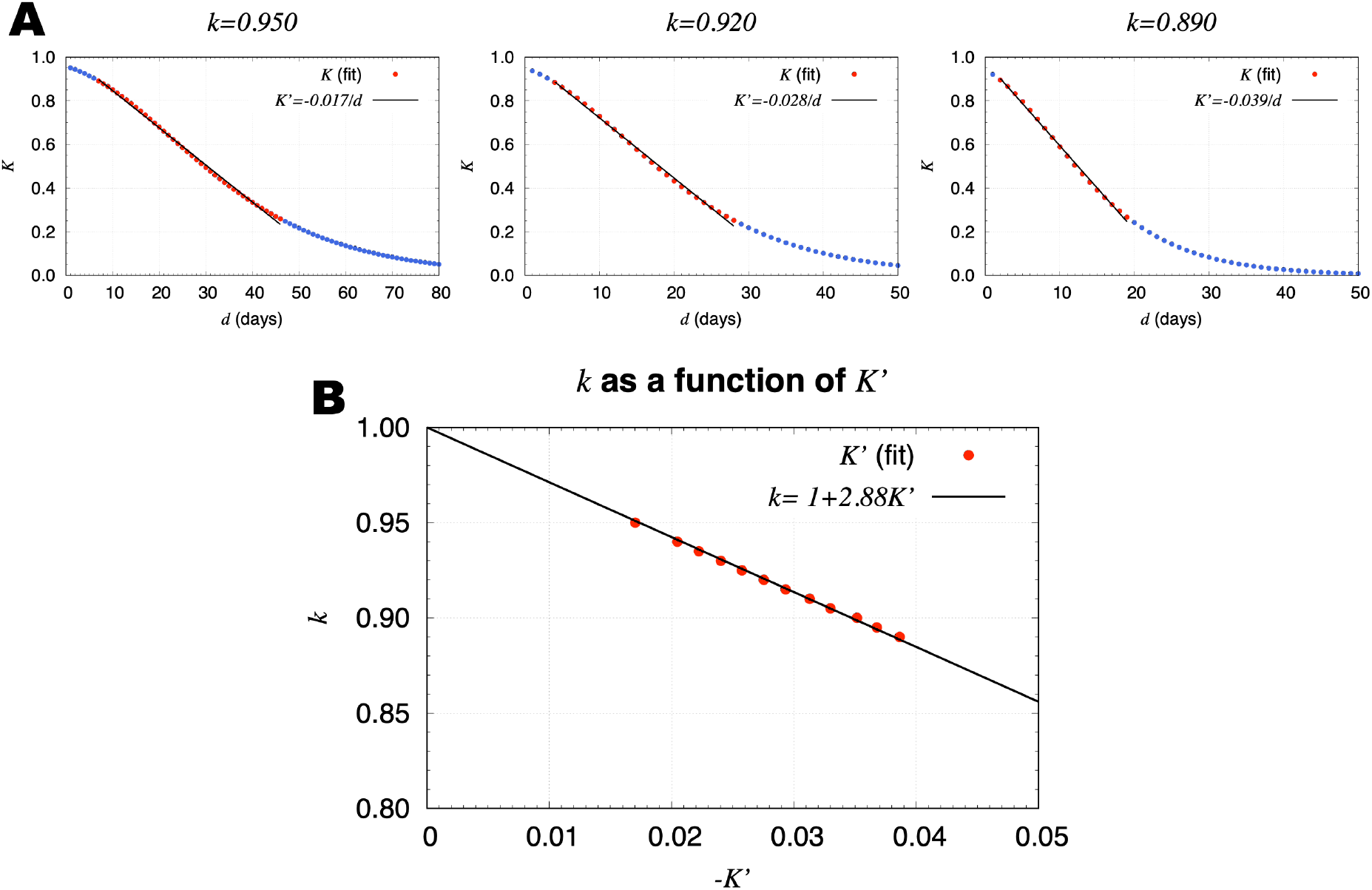
Mock data of the *K* value and *K*′ dependence of *k*. (**A**) The dumping constant *k* was assumed to be 0.95 (**left**), 0.920 (**middle**), and 0.890 (**right**). (**B**) *k* as a linear function of *K′* with a constraint of *k*(0) = 1.

**Fig. S2:**
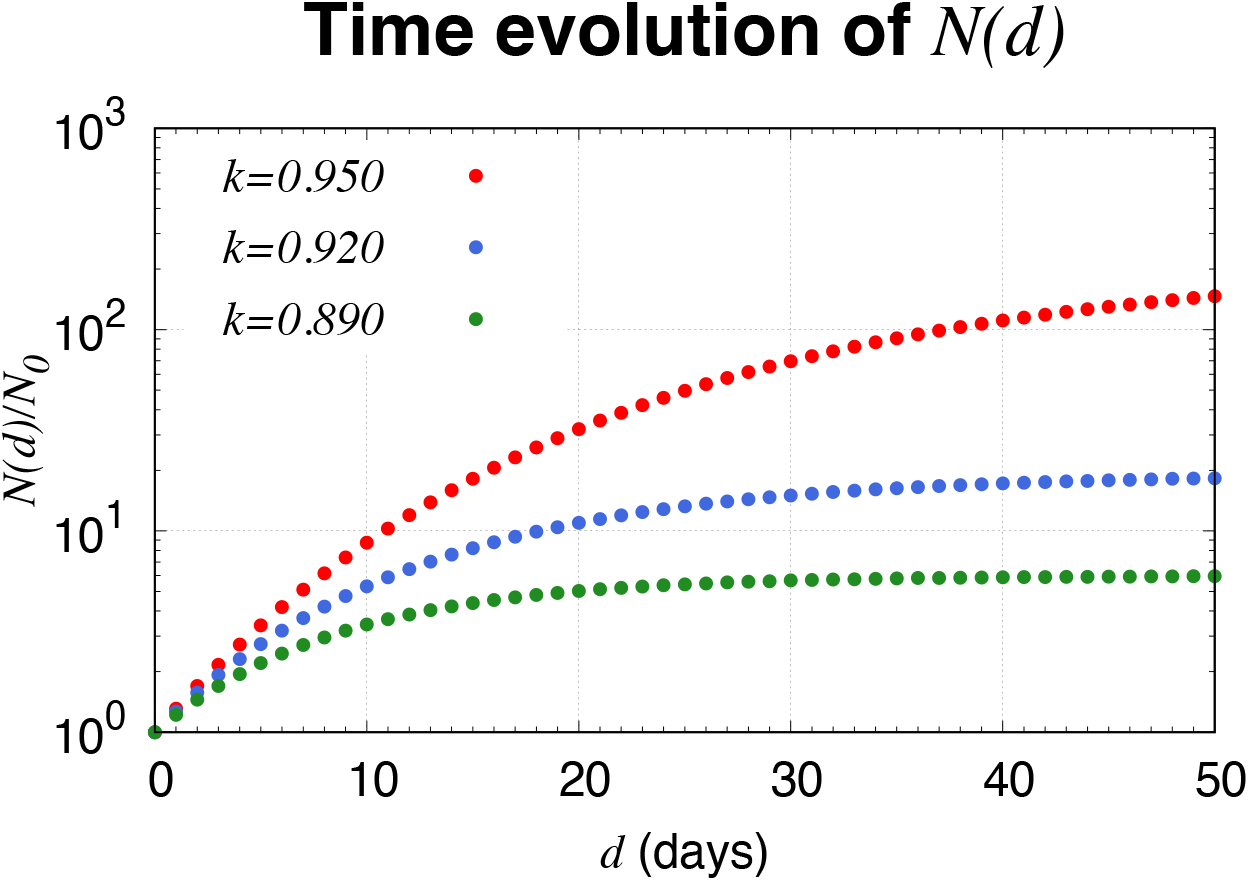
Time evolution of *N*(*d*). Time evolution of *N*(*d*) for the cases with *k* = 0.950, 0.920, and 0.890 (*K′* = −0.017/*d*, −0.028/*d*, and −0.039/*d*) normalized to *N*_0_ by setting *K* = 0.9 on the day zero.

## References

1. Coronavirus Source Data. https://ourworldindata.org/coronavirus-source-data; see also, M. Idogawa, S. Tange, H. Nakase and T. Tokino, “Interactive web-based graphs of novel coronavirus COVID-19 cases and deaths per population by country. Clinical Infectious Diseases” https://dx.doi.org/10.1093/cid/ciaa500

2. The COVID Tracking Project, https://covidtracking.com/data/us-daily

3. Nippon Television Network Corporation, COVID-19 special site. https://www.news24.jp/archives/corona_map/index2.html

4. H. W. Hethcote, “Qualitative analysis of communicable disease models” Math. Biosci., 28, 335–356 (1976).

5. G. Sala, T. Miyakawa “Association of BCG vaccination policy with prevalence and mortality of COVID-19” MedRxiv 2020.03.30.20048165, 6 April 2020. https://doi.org/10.1101/2020.03.30.20048165 (2020).

6. A. Iwasaki, N. D. Grubaugh, “Why does Japan have so few cases of COVID19?” EMBO Mol Med (2020) 0. https://doi.org/10.15252/emmm.202012481 (2020).

